# Unveiling the gap of heart failure: a DATASUS study

**DOI:** 10.1101/2024.06.16.24308996

**Authors:** Vivian Cardoso Batista, Renato Lima Vitorasso, Vicky Nogueira-Pileggi, Renato Mantelli Picoli, Elizabeth Bilevicius

## Abstract

Heart failure (HF) decompensation is the main cause of hospitalizations in developed countries. In Brazil, it represents the third general cause. Aiming to analyze the treatment journey for different types of HF in Brazil, the present study seeks to define a flowchart and clinical rationale that covers the procedures (and their respective frequency) in patients with HF in the Brazilian Unified Health System (SUS) included in DATASUS database. By doing so, the final objectives were: a) To identify potential patients with HF to present an estimate of the underreporting of the disease in the country; b) To describe the estimated mortality of potential patients with HF in Brazil. We used data from DATASUS, which encompasses information from the Brazilian Unified Health System (SUS). Specifically, we utilized the SUS-SIM (Mortality Information System) and SUS-SIA (Ambulatory Information System). Results: According to the data, we had a potential missing of patients with HF of 54,000 patients per year at diagnosis and 200,000 deaths that could lead to HF (both sexes). Considering the sensitivity analysis when there was a 20% underestimation in the number of potential HF cases, the underestimation rate of cases diagnosed with HF was 12%. We also found that when there was an underestimation of 40% in potential cases of death due to HF, there was an average underestimation of 41% in cases of death diagnosed as HF. the results highlight the importance of accurate diagnosis and a comprehensive approach to identifying potential cases of HF to improve the recording and management of this condition. Underestimation of these cases may have significant implications for public health and clinical management of HF emphasizing the need for strategies to increase early detection and adequate case recording. The next steps would be how much this underestimation impacts the public health in Brazil, particularly in terms of financial resources.

## Introduction

According to the World Health Organization (WHO), cardiovascular diseases (CVD) are a group of diseases whose majority can be prevented through a behavioral approach, which represent the main cause of deaths worldwide. It is estimated that 17.9 million people in 2019 died from CVD, representing 32% of all deaths globally(1).

Heart failure (HF) stands out as a highly prevalent condition among CVDs, presenting high morbimortality rates. It has been estimated that HF alone affected 23 million people in 2017, with a projected increase of around 46% by 2030 (3). HF is defined as a clinical syndrome with symptoms and/or signs caused by structural and/or functional cardiac abnormality and corroborated by elevated natriuretic peptides and/or objective evidence of pulmonary or systemic congestion(2).

In Brazil, between 2008 and 2018, HF was the main cause of hospitalizations for CVD with more than 2 million hospitalizations having been recorded and more than 252 thousand deaths. The risk of developing HF is directly proportional to aging. Even though the prevalence is higher among elderly, an increase in prevalence of this condition in the 50-year-old group is being observed. This high hospitalization and mortality rates were responsible for health services expenses that exceeds 3 billion Brazilian Reais (626.5 million US Dollars) only in 2019(3).

HF decompensation is the main cause of hospitalizations in developed countries. In Brazil it represents the third general cause (4). Furthermore, it is believed that there is a high rate of underdiagnosis due to the presence of other comorbid diseases that can be diagnostic confounding factors, especially in cases were HF presents with mild symptoms.

According to the Brazilian Chronic Heart Failure Treatment Guideline, the patient’s prognosis does not depend on the HF subtype and is worse compared to the general population(5).Even though it was observed a decrease in hospitalizations due to HF during the last decade, the number of deaths remained stable. It is important to highlight that mortality rates are possibly underestimated since more than 50% of HF patients experience sudden death. (6).

Aiming to analyze the treatment journey for different types of HF in Brazil, the present study seeks to define a flowchart and clinical rationale that covers the procedures (and their respective frequency) in patients with HF in the Brazilian Unified Health System (SUS) included in DATASUS database. By doing so, the final objectives were:

a. To identify potential patients with HF to present an estimate of the underreporting of the disease in the country.
b. To describe the estimated mortality of potential patients with HF in Brazil.

## Materials and Methods

We sought to understand in the literature the patient journey in Brazil. In this study, we employed data from DATASUS, which encompasses information from the Brazilian Unified Health System (SUS). Specifically, we utilized the SUS-SIM (Mortality Information System) and SUS-SIA (Ambulatory Information System). The data from these systems are accessible to the public via FTP in .*dbc* format, and the SIA-SUS data were converted to .*parquet*. Both data extraction and variable calculations were conducted in a computational environment using R (version 4.2.0) and Python (version 3.10.6). There was no need to seek the approval of an institutional review board before the study began. As previously stated, all the human participants information utilized in this study was obtained through the DATASUS database, a publicly available dataset of the public health system in Brazil. Furthermore, All the data contained in the DATASUS database is fully anonymized when it comes to patients’ personal information. Therefore, no ethical approval or consent was needed. The data utilized was accessed and extracted on September 12^th^ 2023. In this section, we present flowcharts delineating the inclusion criteria for the subgroups under study. This was elaborated according to cardiology specialists regarding patient journey.

The inclusion criteria varied according to the group to be studied in the present project.

The study interval of the present work is highlighted as being from 2018 to 2022 (However, since the patient may have had their HF record or potential HF classification prior to 2018, 3 previous years were included in the extraction to obtain the cut-off without a bias towards the classification of the first years).

1. All those who had at least one record of ICD I50 in any APAC (Ambulatory Procedure Authorization) of DataSUS-SIA were classified as patients with HF.
2. all those deaths that had an ICD I50 note on the death certificate in the base cause, or lines A, B, C, D and II, were classified as HF-related deaths.
3. all those who complied with the flowchart in Fig. 1 and did not have an ICD I50 record were classified as potential patients with HF.
4. Finally, all those who were marked as such in Fig. 2 and did not have an ICD I50 record on the certificate were identified as potential deaths related to HF. It is worth noting that the ICDs indicated in the figure above was searched in all available fields on the death certificate, except for the underlying cause. Additionally, the cause of death had the following subclassifications:

**Fig 1.**
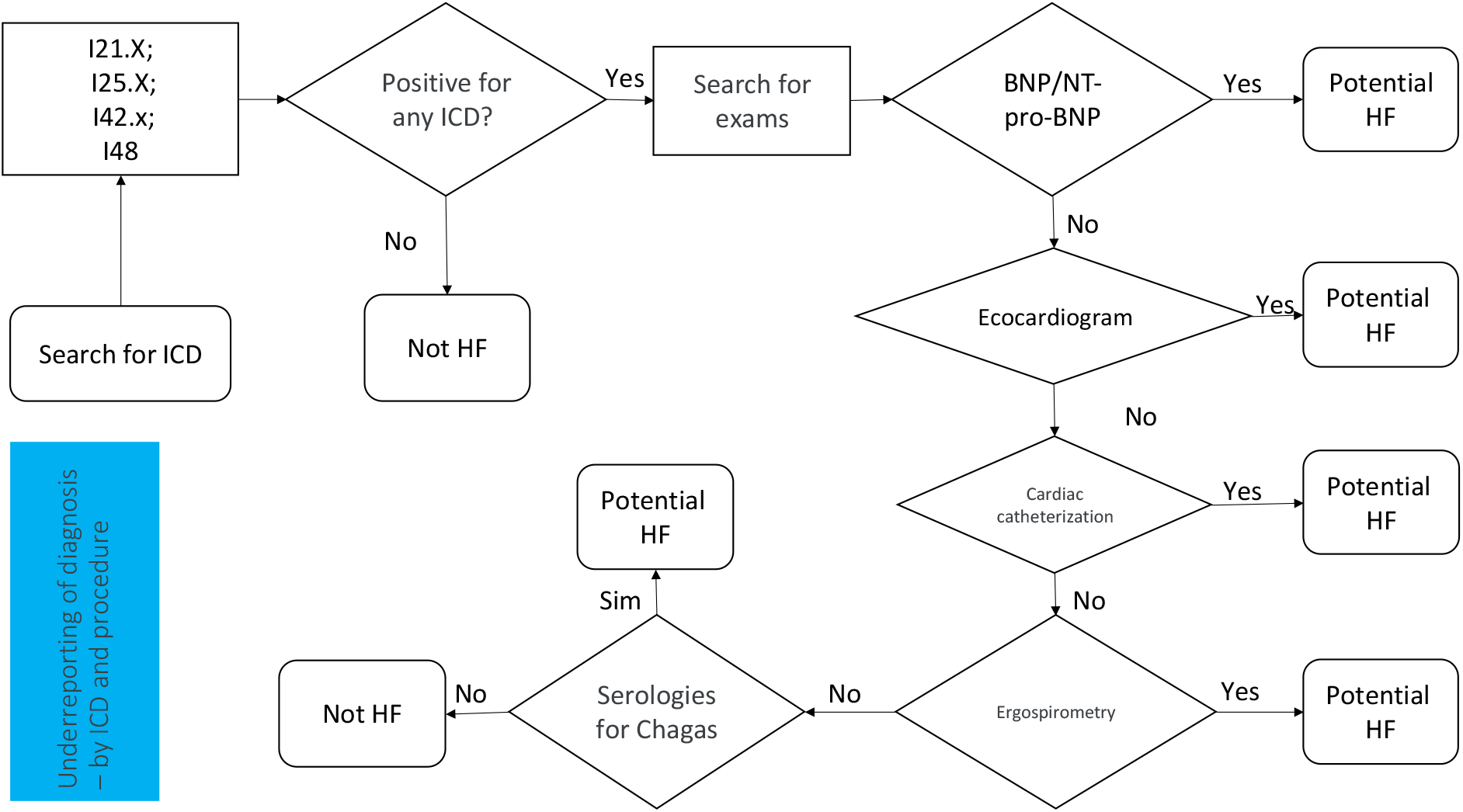
Flowchart to be carried out to find underreported patients by ICD and procedure

**Fig 2.**
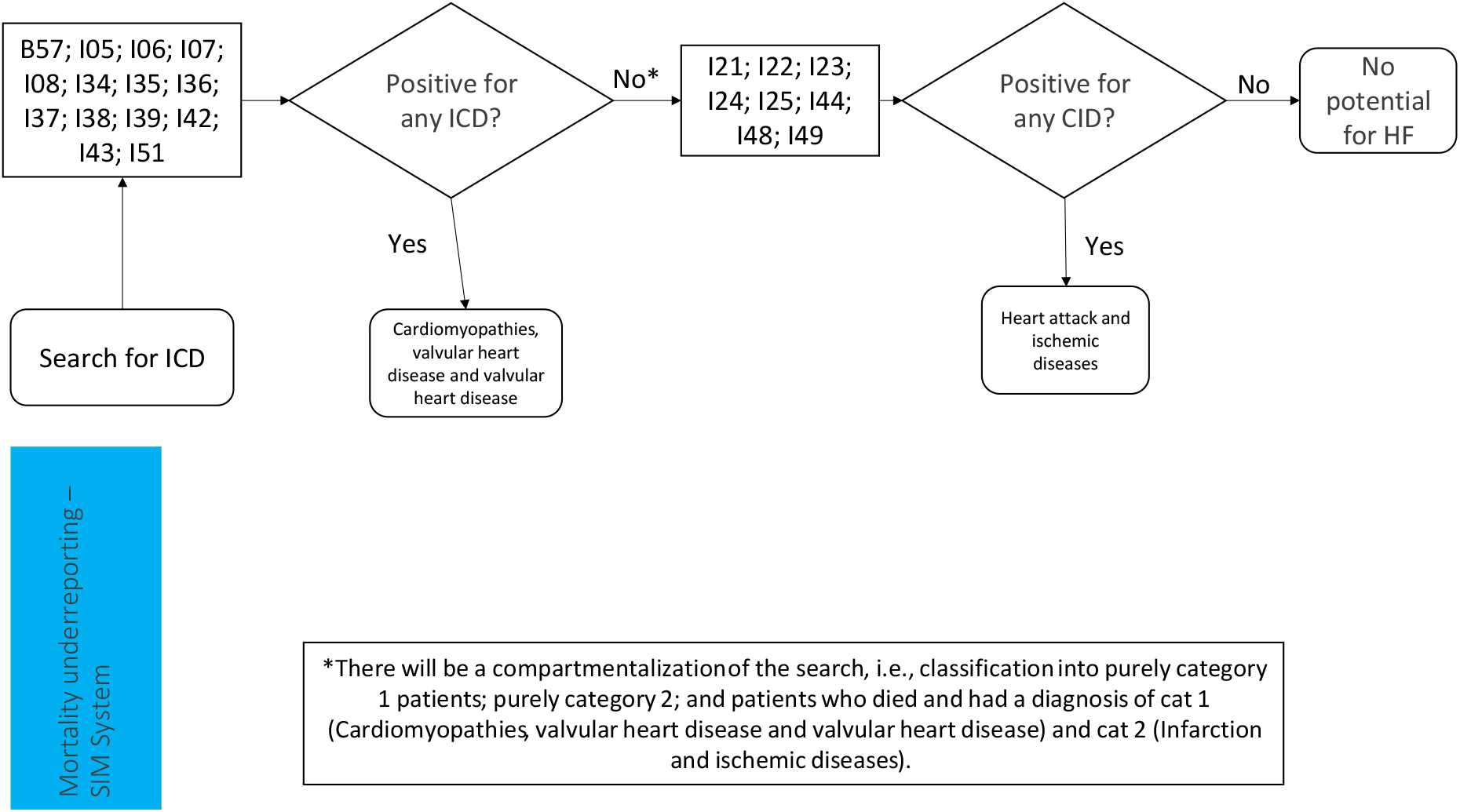
Flowchart to find underreported patients regarding mortality to be carried out in the next step. HF = heart failure. HF = heart failure

a. Cardiomyopathies, valvular heart disease, valvular heart disease.
b. Heart attack and ischemic diseases.

## Sensitivity Analysis

We use as Underestimation rate the following equation:

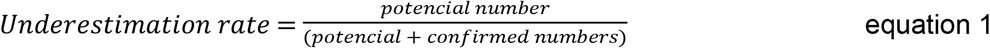

where this rate was applied for both cases and number of deaths. The deterministic sensitivity analysis was conducted to assess the rate of underestimation in A) ambulatorial diagnoses and B) deaths related to heart failure (HF) over the studied time interval. This analysis involves considering values ranging from 0% to 100% (0%, 20%, 40%, 60%, 80%, 100%) as the proportion of potential patients or deaths that are correctly identified as actual patients or deaths related to HF. For instance, when determining the underestimation rate for deaths, assuming 20% of potential patients are correctly identified as actual heart failure (HF) patients, the calculation follows this equation:

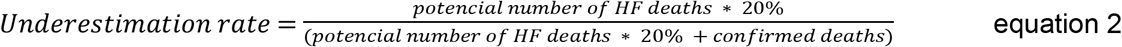

This approach facilitates a thorough investigation into the influence of different proportions of potential patients accurately classified as HF patients on underestimation rates.

## Results

### Population characteristics

Table 1 shows the description of the population separated by patients with a diagnosis of HF and potential HF patients during the years of 2018 to 2022. Table 2 show the description of patients confirmed deaths by HF and those with other types that are potentially HF patients.

**Table 1.** Description of the population of confirmed patients and potential cases of HF.

|  | Age at diagnosis (years) | Male n (%) | Patients |
| --- | --- | --- | --- |
| Diagnosed |  |  |  |
| 2018 | 57.25 ± 21.35 | 32,674.00 (45.19) | 72,309 |
| 2019 | 57.83 ± 21.33 | 35,637.00 (46.02) | 77,445 |
| 2020 | 59.86 ± 19.75 | 30,663.00 (47.98) | 63,905 |
| 2021 | 60.26 ± 19.16 | 44,431.00 (48.91) | 90,842 |
| 2022 | 61.15 ± 18.18 | 62,291.00 (50.58) | 123,155 |
| Potential HF |  |  |  |
| 2018 | 61.02 ± 14.55 | 26,462.00 (53.74) | 49,242 |
| 2019 | 61.94 ± 14.06 | 28,810.00 (54.71) | 52,659 |
| 2020 | 62.45 ± 13.41 | 23,556.00 (55.29) | 42,603 |
| 2021 | 62.48 ± 13.96 | 28,139.00 (55.19) | 50,986 |
| 2022 | 62.66 ± 14.49 | 28,136.00 (54.79) | 51,351 |

**Table 2.** Description of confirmed patient deaths and potential HF-related deaths.

|  | Age on day of death (years) | Male n (%) | Number of deaths |
| --- | --- | --- | --- |
| Diagnosed |  |  |  |
| 2018 | 74.68 ± 17.42 | 53,239.00 (49.10) | 108,430 |
| 2019 | 74.88 ± 17.14 | 53,935.00 (49.14) | 109,758 |
| 2020 | 74.79 ± 15.98 | 58,719.00 (49.83) | 117,849 |
| 2021 | 74.88 ± 16.02 | 65,535.00 (49.65) | 131,981 |
| 2022 | 75.36 ± 16.41 | 64,342.00 (48.96) | 131,411 |
| Potential HF |  |  |  |
| 2018 | 70.75 ± 21.57 | 105,724.00 (56.28) | 187,856 |
| 2019 | 70.97 ± 21.16 | 107,688.00 (56.20) | 191,612 |
| 2020 | 71.30 ± 19.20 | 110,659.00 (57.01) | 194,111 |
| 2021 | 71.12 ± 19.07 | 120,797.00 (56.55) | 213,606 |
| 2022 | 71.47 ± 19.52 | 113,179.00 (56.19) | 201,418 |

Fig 3 shows the absolute number of deaths related to potential cases of HF, stratified by “proxy” ICDs, specifically cardiomyopathies and heart attacks and ischemia, which are conditions that make up potential cases of HF. From an analysis of the figure, it is evident that most deaths associated with potential HF cases are attributed to cases of heart attacks and ischemia, representing approximately 180,000 to 200,000 cases, which is equivalent to approximately 98% of potential HF cases. Furthermore, it is possible to observe a slight increase in the number of deaths from the year 2021 onwards.

**Fig. 3.**
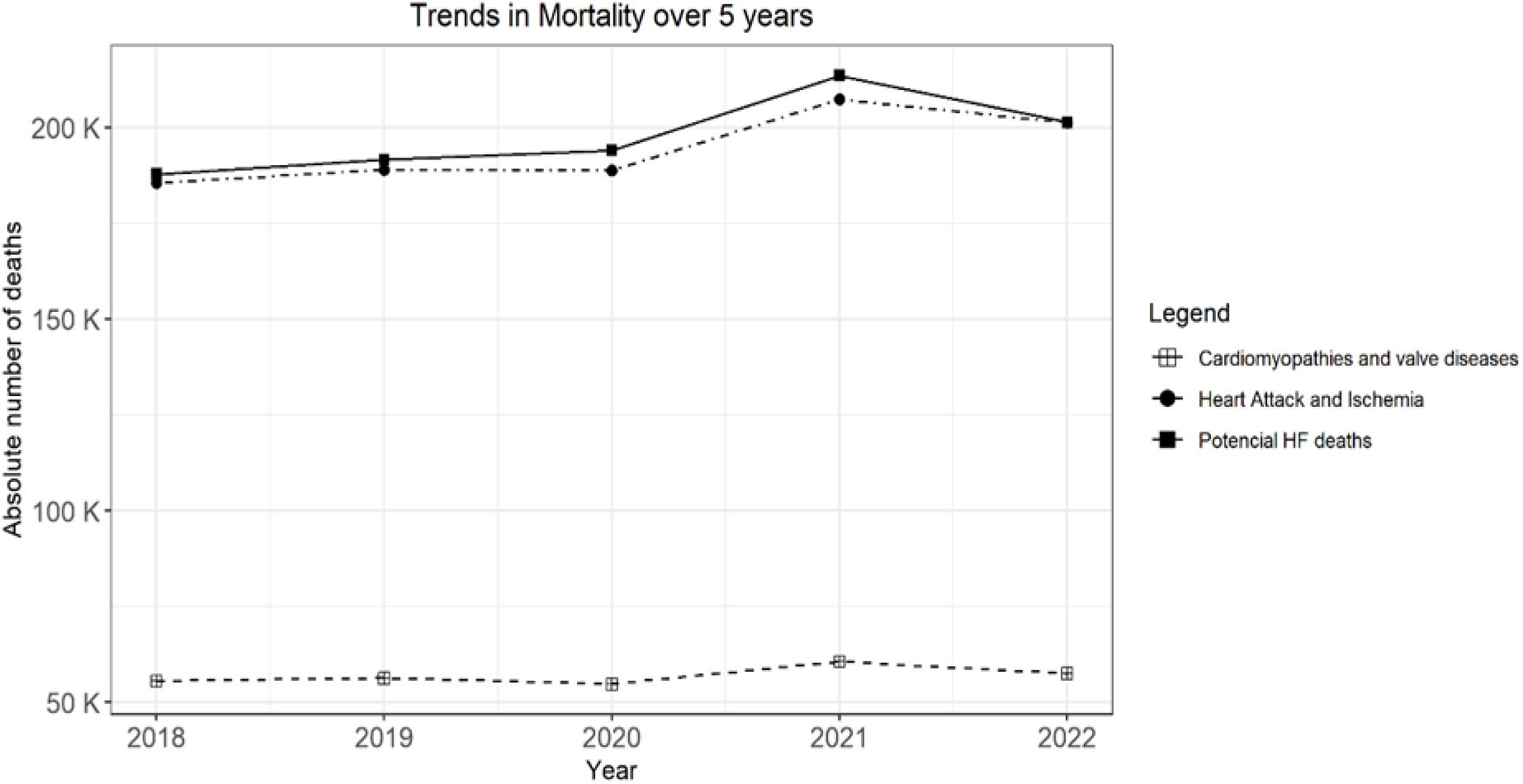
Total number of deaths of patients classified as potential patients with HF divided into the following groups: 1) Cardiomyopathy, valvular heart disease and valvular heart disease; and 2) Ischemia and heart attacks.

Fig 4A presents the representation of the percentage of underestimation of the ambulatorial patients, while Fig 4B shows the percentage of underestimation of the number of deaths of these patients during the 5-year period. In Fig 4A, it is notable that when there was a 20% underestimation in the number of potential HF cases, the underestimation rate of cases diagnosed with HF was 12%. To illustrate another example, when there was an underestimation of 80% in potential HF cases, the underestimation of the number of cases diagnosed with HF reached 35%. Variations were observed from the year 2021 onwards. Similar results can be seen in Fig 4B in relation to deaths due to HF. When there was an underestimation of 40% in potential cases of death due to HF, there was an average underestimation of 41% in cases of death diagnosed as HF. In the case of all potential patients deaths were actually HF cases, approximately 63% of the number of deaths diagnosed as HF were also underestimated.

**Fig 4.**
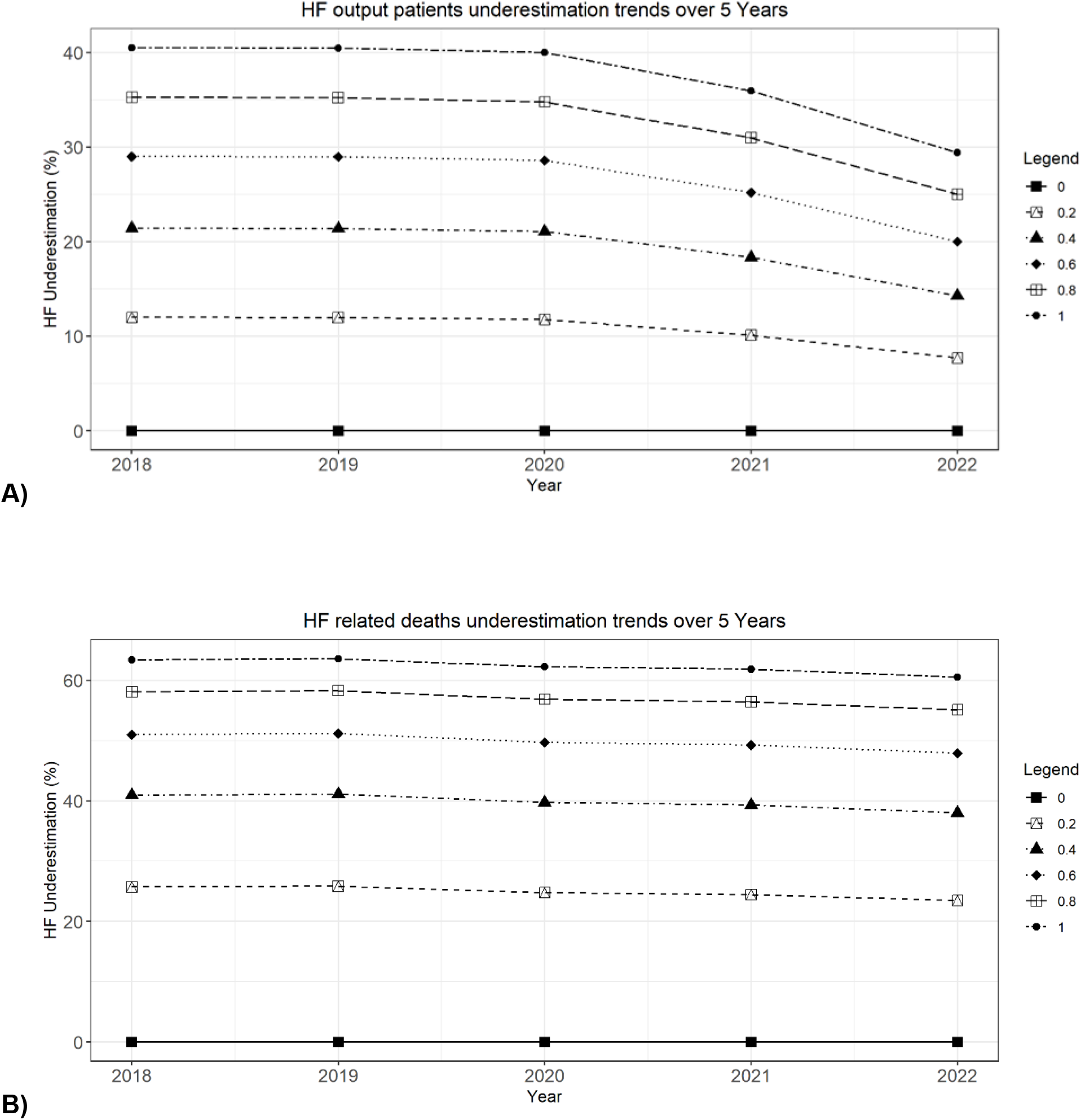
Sensitivity analysis of the rate of underestimation of A) ambulatorial diagnosis and B) deaths related to HF over the time interval studied. The sensitivity analysis will be carried out considering values from 0% to 100% (0; 20; 40; 60; 80; 100%) as the proportion of potential patients/deaths as actual patients/deaths related to HF. Example, to calculate the underestimation rate of deaths at 20%: (potential number of deaths related to HF * 20%) / (potential number of deaths related to HF * 20% + number of deaths with I50 registration).

## Discussion

To the best of our knowledge, this is the first article using public database of Brazil to hypothesize about the misdiagnosis of heart failure and the mortality burden of this disease. It is well known that HF is a common syndrome that represents the end stage of several heart diseases. Decompensation is the main cause of hospitalizations in developed countries and in Brazil it is the third general cause of hospitalization(4). With the strong possibility that many cases of hospitalizations due to HF are underreported because they are associated with other pathologies especially in cases were HF presents with mild symptoms, we create a flowchart to understand this phenomenon.

In our study, the cases of HF were similar between men and women, with a slightly inclination for cases of men in the potential cases. We also found an increase on number of deaths in the studied timeline. This phenomenon could be because aging is a significant factor in the development and progression of heart failure, with the risk increasing as individuals grow older(7) or due to increased number of diagnosis of the disease(8). It is important to stat that the COVID-19 during 2020 contributed to lower numbers in all diseases around the world including HF. The decrease in hospitalizations for non-COVID-19 reasons appears to be linked to hospital overcrowding during periods of COVID-19 surges, coupled with reduced demand for healthcare from individuals concerned about contracting the virus(9).

In 2016 a systematic review analyzed the burden of HF in Latin America countries (10). The study included 143 studies of which 64% were in Brazil, pointing to an incidence of the disease of 199 cases per 100,000 persons-years .The mean percentage of men was 61.07% ±11.48% with a mean age of 60.34± 8.98 years(10). Our study showed that the actual diagnose of HF was in similar age for both HF and potential (around 59 years and 61 years, respectively) with most men in the potential and equal in actual patients with HF.

It is important to emphasize that our study used data from ambulatorial patients, and our study did not show the prevalence of the disease once the patients who seek primary care in the public health care system were the ones studied. This gives us reason to infer that the patients flagged as potential HF cases were in fact cases of HF. That was also the reason we performed a sensitivity analysis that showed that even smaller number of 20% as potential HF, the underestimation of actual ambulatorial cases is 12%.

A study published in 2021 by Wong et al (11) have already highlighted to the relevance of HF misdiagnosis (11). In the review, misdiagnosis ranges from 16.1% in the case of patients discharged from hospital with a diagnosis of HF to 68.5% from general practitioner referrals for HF who do not have left ventricular dysfunction, valvular heart disease, or atrial fibrillation (11). These findings suggests that one of the main reasons for misdiagnosis could be the presence of less severe symptoms. This was the same assumption of our study.

HF is frequently misdiagnosed due to several factors inherent in its presentation and clinical assessment(12). Firstly, HF symptoms such as shortness of breath, fatigue, and edema can overlap with those of other cardiovascular and respiratory conditions, leading to diagnostic ambiguity(13). Moreover, the clinical manifestations of HF may vary widely among patients, making it challenging to recognize consistently across different individuals. Additionally, healthcare providers may encounter difficulties in interpreting diagnostic tests accurately, especially in cases where imaging and biomarkers yield inconclusive results or where comorbid conditions complicate the diagnostic process(14,15). Ultimately, the misdiagnosis of HF underscores the importance of enhanced clinical awareness, comprehensive evaluation strategies, and continued medical education to improve diagnostic accuracy and optimize patient outcomes. Our data corroborate with this potential HF underestimation diagnosis.

Regarding mortality, we represent in Figure 3 the total of deaths of potential patients with HF. Also in the sensitivity analysis we reported that if there was an underestimation of 40% in potential cases of death due to HF, there was an average underestimation of 41% in cases of death diagnosed as HF. Additionally, we represent the number of deaths in subgroups of diseases such as heart attack and ischemia and cardiomyopathies, showing that there is a predominance in the first subgroup over the second. Almost all deaths flagged as potential HF are from patients with heart attack and ischemia in their main cause. It is well known that heart failure often accompanies other cardiovascular conditions, compounding the risk of adverse outcomes. Early diagnosis, comprehensive management strategies, adherence to treatment plans, and lifestyle modifications play pivotal roles in reducing mortality rates and improving survival outcomes among individuals living with heart failure.

The data of the present study point to an overall increase in HF-related ambulatorial patients and mortality and potential HF cases over five years, with some notable fluctuations and significant variations in specific years. Furthermore, the underestimation of potential HF cases potentially impacted the underestimation of diagnosed cases and deaths from HF, highlighting the importance of an accurate assessment of these data for the effective management of this health condition.

## Conclusions

In summary, the results highlight the importance of accurate diagnosis and a comprehensive approach to identifying potential cases of IC to improve the recording and management of this condition. Underestimation of these cases may have significant implications for public health and clinical management of IC, emphasizing the need for strategies to increase early detection and adequate case recording. The next steps would be how much this underestimation impacts the public health in Brazil, particularly in terms of financial resources.

## Data Availability

All files are available from the DATASUS database. Available at: https://datasus.saude.gov.br/

## References

1. World Health Organization. Cardiovascular diseases (CVDs) [Internet]. 2021. Disponível em: https://www.who.int/news-room/fact-sheets/detail/cardiovascular-diseases-(cvds)

2. Bozkurt B, Coats AJ, Tsutsui H, Abdelhamid M, Adamopoulos S, Albert N, et al. Universal Definition and Classification of Heart Failure. J Card Fail. abril de 2021;27(4):387–413.

3. Arruda VL de, Machado LMG, Lima JC, Silva PR de S. Trends in mortality from heart failure in Brazil: 1998 to 2019. Rev Bras Epidemiol Braz J Epidemiol. 2022;25:E220021.

4. Bocchi EA, Vilas-Boas F, Perrone S, Caamaño AG, Clausell N, Moreira M da CV, et al. I Diretriz Latino-Americana para avaliação e conduta na insuficiência cardíaca descompensada. Arq Bras Cardiol [Internet]. setembro de 2005 [citado 3 de março de 2023];85. Disponível em: http://www.scielo.br/scielo.php?script=sci_arttext&pid=S0066-782X2005002200001&lng=pt&nrm=iso&tlng=pt

5. Rohde LEP, Montera MW, Bocchi EA, Clausell NO, Albuquerque DC de, Rassi S, et al. Diretriz Brasileira de Insuficiência Cardíaca Crônica e Aguda. Arq Bras Cardiol [Internet]. 2018 [citado 1o de março de 2023]; Disponível em: https://www.scielo.br/scielo.php?script=sci_arttext&pid=S0066-782X2018001500436

6. Alexsander R, De Las Casas Bessa L, Dias Silveira AV, Gualberto Souza I, Silveira Ferreira GF, Pacheco Souza G, et al. Análise Epidemiológica por Insuficiência Cardíaca no Brasil. Braz Med Stud [Internet]. 12 de abril de 2022 [citado 27 de fevereiro de 2023];6(9). Disponível em: https://bms.ifmsabrazil.org/index.php/bms/article/view/224

7. Strait JB, Lakatta EG. Aging-associated cardiovascular changes and their relationship to heart failure. Heart Fail Clin. janeiro de 2012;8(1):143–64.

8. Eriksson H. Heart failure: a growing public health problem. J Intern Med. fevereiro de 1995;237(2):135–41.

9. Menezes-Filho N, Komatsu BK, Villares L. The impacts of COVID-19 hospitalizations on non-COVID-19 deaths and hospitalizations: A panel data analysis using Brazilian municipalities. PLOS ONE. 14 de dezembro de 2023;18(12):e0295572.

10. Ciapponi A, Alcaraz A, Calderón M, Matta MG, Chaparro M, Soto N, et al. Burden of Heart Failure in Latin America: A Systematic Review and Meta-analysis. Rev Espanola Cardiol Engl Ed. novembro de 2016;69(11):1051–60.

11. Wong CW, Tafuro J, Azam Z, Satchithananda D, Duckett S, Barker D, et al. Misdiagnosis of Heart Failure: A Systematic Review of the Literature. J Card Fail. setembro de 2021;27(9):925–33.

12. Bottle A, Kim D, Aylin P, Cowie MR, Majeed A, Hayhoe B. Routes to diagnosis of heart failure: observational study using linked data in England. Heart Br Card Soc. abril de 2018;104(7):600–5.

13. van Riet EES, Hoes AW, Limburg A, Landman MAJ, van der Hoeven H, Rutten FH. Prevalence of unrecognized heart failure in older persons with shortness of breath on exertion. Eur J Heart Fail. julho de 2014;16(7):772–7.

14. Mant J, Doust J, Roalfe A, Barton P, Cowie MR, Glasziou P, et al. Systematic review and individual patient data meta-analysis of diagnosis of heart failure, with modelling of implications of different diagnostic strategies in primary care. Health Technol Assess Winch Engl. julho de 2009;13(32):1–207, iii.

15. Barents M, van der Horst ICC, Voors AA, Hillege JL, Muskiet F a. J, de Jongste MJL. Prevalence and misdiagnosis of chronic heart failure in nursing home residents: the role of B-type natriuretic peptides. Neth Heart J Mon J Neth Soc Cardiol Neth Heart Found. abril de 2008;16(4):123–8.

